# SARS-CoV-2 reinfection with Omicron BA.2.75 subvariants in Thai Adults in Thailand

**DOI:** 10.1101/2023.11.24.23298841

**Authors:** Suvichada Assawakosri, Natthinee Sudhinaraset, Jira Chansaenroj, Nungruthai Suntronwong, Sitthichai Kanokudom, Natach Nalinpakorn, Napat Tantipraphat, Amica Sethabutra, Sittisak Honsawek, Yong Poovorawan

## Abstract

Omicron subvariants of SARS-CoV-2 may resist vaccine- or infection-induced immunity thereby increasing the risk of reinfections in previously infected persons. This study aimed to investigate the clinical severity and the average time to the onset of Omicron reinfection. This survey study collected clinical data on Omicron reinfection. Information on time of infection, reinfection interval, overall clinical presentation, and severity of infection was reported. The total prevalence of symptoms among 201 participants was significantly higher in the first infection (risk difference (RD), 9.86%; 95% CI, 7.54–12.19]) compared to the second infection, and the hospitalization rate among all participants was significantly lower for the second infection than the primary infection (odds ratio (OR), 6.25; 95% CI, 2.158−24.71). The prevalence of symptoms compared with the first infection with pre-Omicron variants was similar to that of the first infection with the Omicron variant (RD, 2.56%; 95% CI, -6.14–1.01). However, the hospitalization rate for pre-Omicron primary infection was significantly higher (OR, 6.76; 95% CI, 2.87–15.87]) than that observed with Omicron variants. The severity of the primary infection and of a pre-Omicron variant was greater than that of a secondary infection or with an Omicron variant.

## Introduction

In March 2020 the World Health Organization (WHO) declared Coronavirus disease (COVID-19) a worldwide pandemic. Since then, there have been over 676 million cases and 6.8 million deaths worldwide. As of January 2023, more than 4.7 million cases and 33 thousand deaths have been reported in Thailand. In the current pandemic stage, new SARS-CoV-2 Omicron variants with a high ability to escape vaccine- and infection-induced immunity have raised the concern of potential reinfection among previously infected individuals.

In August 2020, the first reinfection case of COVID-19 was documented in Hong Kong [1]. Reinfection of COVID-19 is defined as a person infected with a genetically different variant from the previous infection or probable reinfection after a more than 90-day interval between two infections [2]. Previous studies have demonstrated that the rate of reinfection with COVID-19 increased from <2.7 % to 11% after the Omicron wave started [3]. Symptoms can range from asymptomatic to severe illness due to Omicron infection. However, most patients recovered within a few weeks after infection.

Infected individuals may experience lower levels of severity on the second episode of infection during the transition between Delta and the Omicron wave [4]. However, there are limited data on the severity of the disease in individuals with repeated infections with the Omicron variant, especially infections with BA.4/5 or BA.2.75. This study aimed to investigate clinical data on the severity disease following reinfection during the Omicron wave in the Thai population.

## Methods

### Study design

This study was a survey study. The study protocol was approved by the Institutional Review Board of the Faculty of Medicine of Chulalongkorn University (IRB numbers 750/65). The study was conducted in accordance with the principles of the Declaration of Helsinki and the Good Clinical Practice Guidelines (ICH-GCP). Informed consent was obtained electronically from all participants before the survey started.

### Study participants

In January 2023, subjects participated in this electronic survey study to report COVID-19 symptoms. Data, including participant characteristics such as sex, age, comorbidity, occupation, COVID-19 vaccination history, and infection history were obtained from all participants. The inclusion criteria were Thai adults aged over 18 years with a history of two COVID-19 infections. The time interval between two episodes of infection was >90 days. Participants with incomplete infection data, who were not immunocompetent, who were receiving immunosuppressive drugs, and participants who were pregnant or breastfeeding were excluded from the study. The timing of the infection was used to assign the pre-Omicron or Omicron infection period. According to the sequenced surveillance data for the predominant SARS-CoV-2 variant obtained in our previous study in Thailand [5], infections that occurred before January 2022 were considered from the pre-Omicron infection period.

### Statistical analysis

All analyses were performed using the Statistical Package for the Social Sciences (SPSS) v.29 (SPSS Inc., Chicago, IL, USA). Comparison of categorical data, including sex and comorbidities, was performed using Pearson’s chi-square test. The Mann–Whitney U test was used to compare between groups (nonparametric analysis). The general risk difference (RD) and 95% confidence intervals (CI) of acute symptomatic infections were estimated. Figures were generated using GraphPad Prism v9.0 (GraphPad Software, San Diego, CA, USA) and RStudio with R statistical software version 4.2.0 (R Project for Statistical Computing).

## RESULTS

### Demographic data

In January 2023, a total of 201 adults were enrolled in this survey study. All participants were Thai adults, could be divided into two groups: 169 non-healthcare workers (non-HCW) and 32 healthcare workers (HCW). The majority of the participants were female, comprising 120 (59.7%) of the 201 participants. The overall mean age was 44.3 [20–81] years, while the mean age (range) in the non-HCW and HCW groups was 45.1 [20–81] and 39.9 [23–66] years, respectively. For the vaccination profile, the participants who received two, three, four or more doses of the COVID-19 vaccine were combined. Most participants completed at least two primary doses (99.0%) and one booster dose (89.5%). Generally, there are no significant differences in the baseline characteristics, including sex, age, common underlying disease, or the timing of infection between the two groups. The general demographics are shown in Table 1.

**Table 1.** Demographics and baseline characteristics of the enrolled participants. Abbreviations: HCW: Health care worker; Non-HCW: non-Health care worker; NA: Not-available; COPD: Chronic obstructive pulmonary disease.

|  | **Total** | **Non-HCW** | **HCW** |
| --- | --- | --- | --- |
| **Total number (n)** | 201 | 169 | 32 |
| **Mean age (range) years** | 44.3 (20–81) | 45.1 (20–81) | 39.9 (23–66) |
| **Sex** |  |  |  |
| Male (%) | 78 (38.8) | 70 (41.4) | 8 (25.0) |
| Female (%) | 120 (59.7) | 97 (57.4) | 23 (71.9) |
| NA (%) | 3 (1.5) | 2 (1.2) | 1 (3.1) |
| **Underlying disease (%)** |  |  |  |
| Hypertension | 30 (14.9) | 25 (14.8) | 5 (15.6) |
| Diabetes Mellitus | 11 (5.5) | 10 (5.9) | 1 (3.1) |
| Cardiovascular diseases | 5 (2.5) | 5 (3.0) | - |
| Obesity | 7 (3.5) | 7 (4.1) | - |
| Dyslipidemia | 28 (13.9) | 23 (13.6) | 5 (15.6) |
| Allergy | 25 (12.4) | 20 (11.8) | 5 (15.6) |
| Asthma | 3 (1.5) | 3 (1.8) | - |
| Thyroid | 5 (2.5) | 4 (2.4) | 1 (3.1) |
| Cancer | 1 (0.5) | 1 (0.6) | - |
| Other (gout, COPD, etc.) | 13 (6.5) | 12 (7.1) | 1 (3.1) |
| **Interval between two infections, (range) days** | 262.8 (96–725) | 263.0 (96–640) | 261.3 (99–725) |
| **Interval between two infections group** |  |  |  |
| 0–180 days (%) | 62 (30.8) | 52 (30.8) | 10 (31.3) |
| > 180 days (%) | 139 (69.2) | 117 (69.2) | 22 (68.7) |
| **Time of the first infection (%)** |  |  |  |
| Pre-Omicron | 65 (32.3) | 55 (32.5) | 10 (31.3) |
| Omicron | 136 (67.7) | 114 (67.5) | 22 (68.7) |
| **Vaccine Dose (%)** |  |  |  |
| 0 | 1 (0.5) | 2 (0.6) | - |
| 1 | 1 (0.5) | 1 (0.6) | - |
| 2 | 19 (9.5) | 19 (11.2) | - |
| 3 | 68 (33.8) | 65 (37.9) | 4 (12.5) |
| 4 | 82 (40.8) | 62 (36.7) | 20 (62.5) |
| 5 | 28 (13.9) | 21 (12.4) | 7 (21.9) |
| ≥ 6 | 2 (1.0) | 1 (0.6) | 1 (3.1) |

### Timing of infection and reinfection interval

All participants were subdivided into two groups according to the timing of the first infection. The first infections ranged from January 2021 to October 2022: 65 infections were reported during the pre-Omicron wave (32.3%) and 136 infections were reported during the predominant Omicron wave (67.7%). Reinfections peaked during the Omicron BA.4/5 and BA.2.75 predominant wave, as shown in Fig. 1. The average time interval between the first infection (circle) and the reinfection (square) was 262.8 [range 96-725] days. For most participants 139/201 (69.2%), the reinfection interval time was greater than 180 days since the primary infection.

**Fig. 1.**
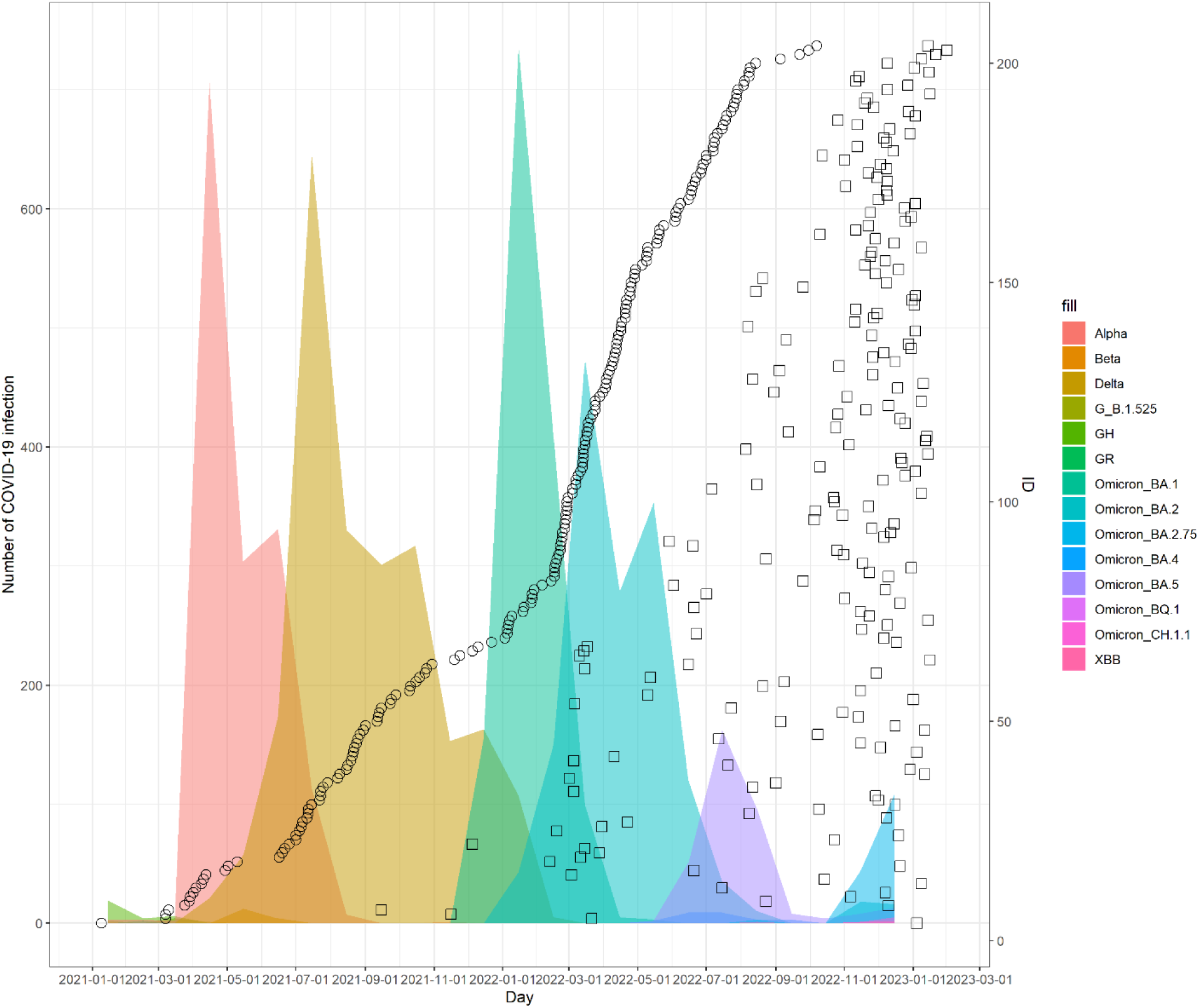
Timeline of COVID-19 reinfection cases. Time course of the first infection (circle) and reinfection (square) were plotted versus the sequencing data of the SARS-CoV-2 predominant variant obtained in our previous study [5].

### Overall clinical presentation and the severity of infection

The prevalence and severity of acute symptoms of the first and second infections were reported. The most common symptoms during the first infection were cough (76.6%), sore throat (73.6%), and runny nose (73.1%), respectively. In comparison, common symptoms during the second episode were runny nose (75.6%), cough (73.6%), and sore throat (64.2%), respectively. Most of the clinical symptoms were presented as mild to moderate in severity. The percentage of severe symptoms was less observed in the second infection compared to the primary exposure to COVID-19 (Supplementary Fig. 1). Statistical analysis showed that the total prevalence of symptoms was significantly higher in the first infection (RD, 9.86% [95% CI; 7.54-12.19]) than in the second infection, as shown in Fig. 2A. However, no significant differences were observed in some common acute symptoms, including runny nose, cough, sputum production, headache, hoarseness, and rash. In summary, these acute symptoms were a common signature clinical symptom of SARS-CoV-2 infection. Overall, the hospitalization rate among all participants was significantly lower for the second infection compared to the primary infection (OR, 6.25 [95% CI, 2.158−24.71]) (Table 2).

**Fig. 2.**
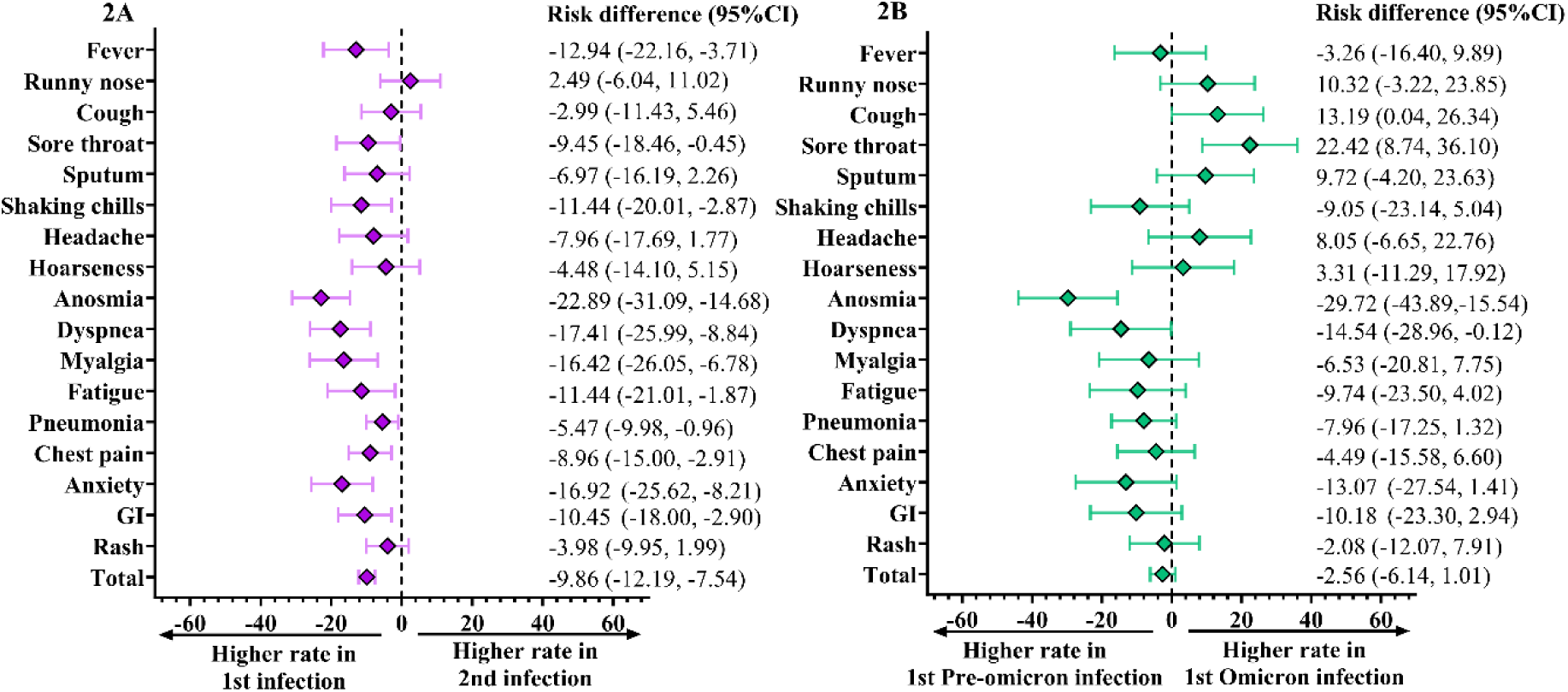
Comparison of the prevalence of acute clinical symptoms. The Forest plot presents the prevalence of any grade of clinical symptom and the comparison of absolute risk differences between two infections (A) between the first and second infection; (B) between the first infection with a pre-Omicron variant and first infection with an Omicron variant.

**Table 2.**
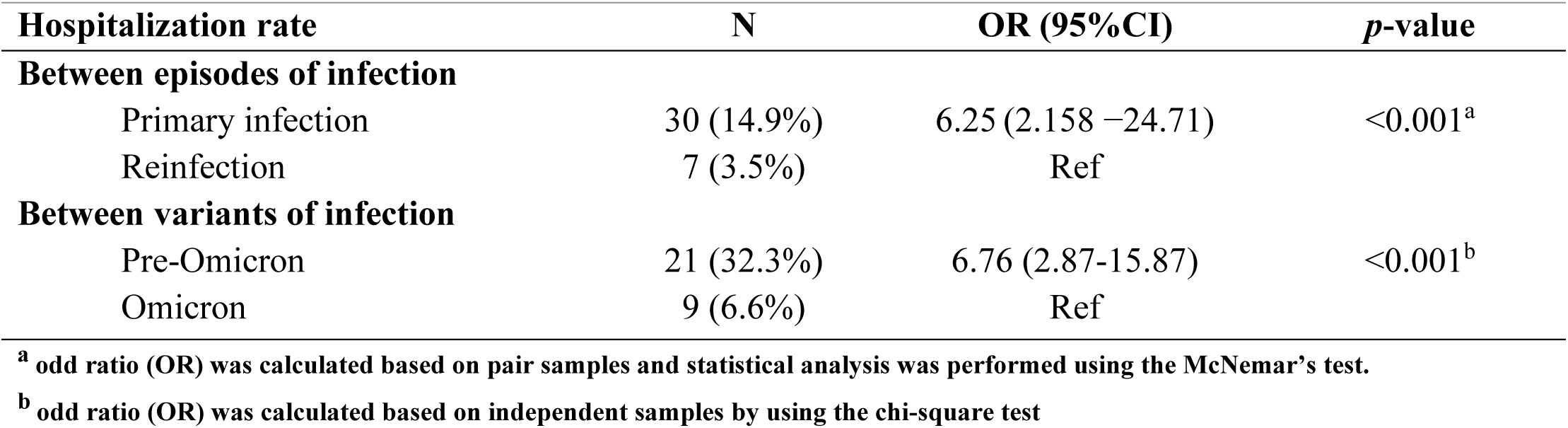
An event of hospitalization compared between two episodes of infection and between variants of primary infection.

| Hospitalization rate | N | OR (95%CI) | *p*-value |
| --- | --- | --- | --- |
| **Between episodes of infection** |  |  |  |
| Primary infection | 30 (14.9%) | 6.25 (2.158 –24.71) | <0.001<sup>a</sup> |
| Reinfection | 7 (3.5%) | Ref |  |
| **Between variants of infection** |  |  |  |
| Pre-Omicron | 21 (32.3%) | 6.76 (2.87-15.87) | <0.001<sup>b</sup> |
| Omicron | 9 (6.6%) | Ref |  |

<sup>a</sup> odd ratio (OR) was calculated based on pair samples and statistical analysis was performed using the McNemar's test.
<sup>b</sup> odd ratio (OR) was calculated based on independent samples by using the chi-square test

### Comparison of the clinical presentation of pre-Omicron and Omicron variant infections

We compared the clinical characteristics between the first infection with those of the different variants and between the two episodes of infection. First, the total prevalence of symptoms between the first infection with the pre-Omicron variants was not significantly different from the first infection with the Omicron variant (RD 2.56% [95% CI; -6.14 to 1.01]) as shown in Fig. 2B. However, the hospitalization rate of the primary infection with pre-Omicron variants was significantly higher (OR, 6.76 [95% CI, 2.87–15.87]) than that observed on infection with Omicron variants (Table 2). Anosmia was significantly higher (RD, 29.7% [95% CI; 15.5–43.9]) in patients with pre-Omicron infection than in the Omicron infection group. At the same time, symptoms of sore throat were significantly higher observed in patients with Omicron infection (RD, 22.4% [95% CI; 8.7, 36.1]).

Comparing the two infection episodes, the group with the first infection with the pre-Omicron variant showed a significantly higher prevalence of lower respiratory symptoms and other systemic symptoms such as anosmia, dyspnea, pneumonia, chest pain, anxiety, myalgia, fatigue, and GI symptoms than that observed on reinfection with the Omicron variant (Fig. 3C). Instead, the group with Omicron primary infection presented a significantly higher prevalence of local upper respiratory symptoms than the repeated Omicron episode, including sore throat, sputum, anosmia, and some other systemic symptoms such as fever, dyspnea, anxiety, myalgia, and fatigue (Fig. 3D). These results suggested that the first infection likely caused more severe disease conditions than the second infection.

**Fig. 3.**
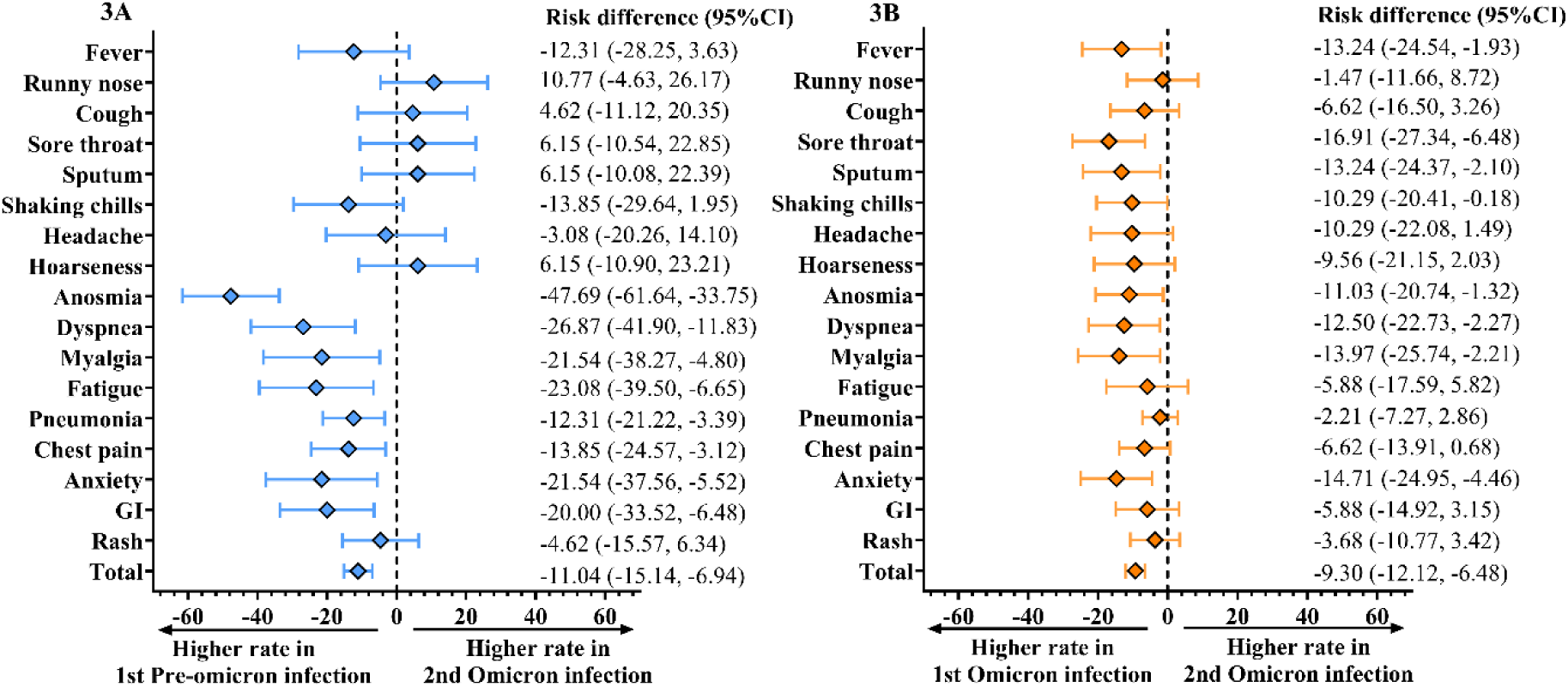
Comparison of the prevalence of acute clinical symptoms in different primary infection strain. The Forest plot presents the prevalence of any grade of clinical symptom and the comparison of absolute risk differences between two infections (A) between the first infection with a pre-Omicron variant and second infection with Omicron variant; and (B) compared between the first infection with an Omicron variant and second infection with Omicron variant.

## Discussion

Following the emergence of several Omicron subvariants, including BA.1, BA.1.1, BA.2, BA.2.12.1, BA.2.75, and BA.4/5, COVID-19 reinfection has increased dramatically worldwide. Due to the waning of antibodies over time after vaccination or natural infection [6] and given the large number of spike mutations harbored by the Omicron variant, patients who were previously infected pre-Omicron variants experienced a second breakthrough infection on exposure to different Omicron subvariants of SARS-CoV-2 [7]. A previous study revealed that the titer of neutralizing antibodies (NAbs) from the Delta-infected serums was significantly lower against the Omicron variant than that of against the Wild-type or Delta variant [8].

Additionally, the accumulation of mutations in spike-RBD among newly emerged Omicron subvariants often contributes to repeated Omicron breakthrough infections [9]. A significant increase in the rate of reinfection in patients with primary Omicron infection (BA.1 or BA.2) after exposure to BA.4 and BA.5 were dominant variants in Qatar [10]. Our study showed that the reinfection events mainly occurred during the introduction of Omicron BA.4/5 and BA.2.75 subvariants in Thailand. Although 70% of the study population had experienced an Omicron primary infection with BA.1 or BA.2, they had repeated Omicron breakthrough infections. Thus, our data demonstrated that the new Omicron subvariant, especially Omicron BA.2.75, could be associated with a higher risk of reinfection among individuals with hybrid immunity.

This study also compared the prevalence of the clinical characteristics and disease features of the two-infection episodes and the reinfection time interval. Our findings showed that the average time to the onset of reinfections was 262 days among all participants. In agreement with a previous study, almost 80% to 90% of immunological memory (both CD4 T cell and B cell memory) of SARS-CoV-2 persisted for 6 to 8 months after primary recovery [11]. Compared to another study from Brazil, our finding showed there was a shorter estimated time interval between two infections (approximately 8- vs. 14-month interval) [12]. Even though protection against reinfection may decrease over time after primary infection, protection against severe symptoms after reinfection was maintained at more than 90% for patients with Omicron infection [13]. Our finding revealed that the prevalence of acute symptoms, the severity of the disease, and the hospitalization rate were significantly lower in the Omicron reinfection episode compared to the primary episode. In support to our results, Sotoodeh et al. also reported that most reinfected cases presented the clinical symptom as asymptomatic or with mild symptoms [14].

Moreover, our study demonstrated that the hospitalization rate in the Omicron primary infection group was significantly lower compared to primary infection with the pre-Omicron variant. Similar to recent studies, the clinical symptoms of Omicron infection were substantially less severe than those of all other variants of SARS-CoV-2 [15, 16]. According to ex *vivo* evidence, the extensive mutation in the spike protein of Omicron altered its replication properties. Since the Omicron variant is highly replicated and accumulates in the upper respiratory tract, it has a lower replication competence in the human lungs [17]. This finding may explain why the Omicron variant does not cause severe lower respiratory symptoms. Another reason for the attenuated symptoms on reinfection might be the level of coverage of vaccination among our study population, since several studies have shown that protection against severe infection was significantly higher in those who completed a booster dose of the COVID-19 mRNA vaccine than in unvaccinated individuals [18].

The primary limitation of our study was the lack of confirmed sequencing analysis of SARS-CoV-2 variants for all participants; thus, we can only attribute the variant strain responsible for infection by relying on the time of infection. Furthermore, a subgroup analyses that stratifies patients by age or vaccination status could not be performed due to the small number of participants. Further study with a larger sample size of participants or the study of estimates rate of SARS-CoV-2 reinfections in the Thai population will contribute to future policy decisions on preventive vaccination strategies.

In summary, our study found that the clinical characteristics of repeated Omicron infections were less severe than those of the primary infection. The estimated interval between the primary and reinfection episode was approximately 8 to 9 months after the primary infection. Furthermore, the viremic effect of the primary Omicron variants was lower than that of infection with the pre-Omicron variant.

## Data Availability

All data produced in the present study are available upon reasonable request to the authors.

## Ethical approval and consent to participate

The study protocol was approved by the Institutional Review Board of the Faculty of Medicine of Chulalongkorn University (IRB numbers 750/65). Electronic informed consent was obtained from all participants before the survey started.

## Author Contributions

Conceptualization, S.A., N.S. and Y.P.; data curation, S.A., N.S. and D.S.; formal analysis, S.A., J.C., N.S. (Nungruthai Suntronwong); methodology, S.A., P.N., N.N., N.T., A.S.; project administration, N.S., S.H. and Y.P.; writing—original draft, S.A.; writing—review and editing, J.C., N.S. (Nungruthai Suntronwong), S.K., and Y.P. All authors have read and agreed to publish this version of the manuscript.

## Acknowledgments

The authors are grateful to the Center of Excellence in Clinical Virology staff, all the participants, BJC Big C Foundation and MK restaurant group public Co., Ltd. for helping and supporting this project.

