## Supplementary figure1 for "SARS-CoV-2 reinfection with Omicron BA.2.75 subvariants in Thai Adults in Thailand": Supplement1.pdf

**Supplementary Figure 1. Comparison of the severity percentage of clinical symptoms between the first (dot fill) and second (solid fill) infection.** The events were classified as mild (no impact on regular activity)/moderate (some limitation of daily activity), and severe (unable to perform or prevented performing daily activities.

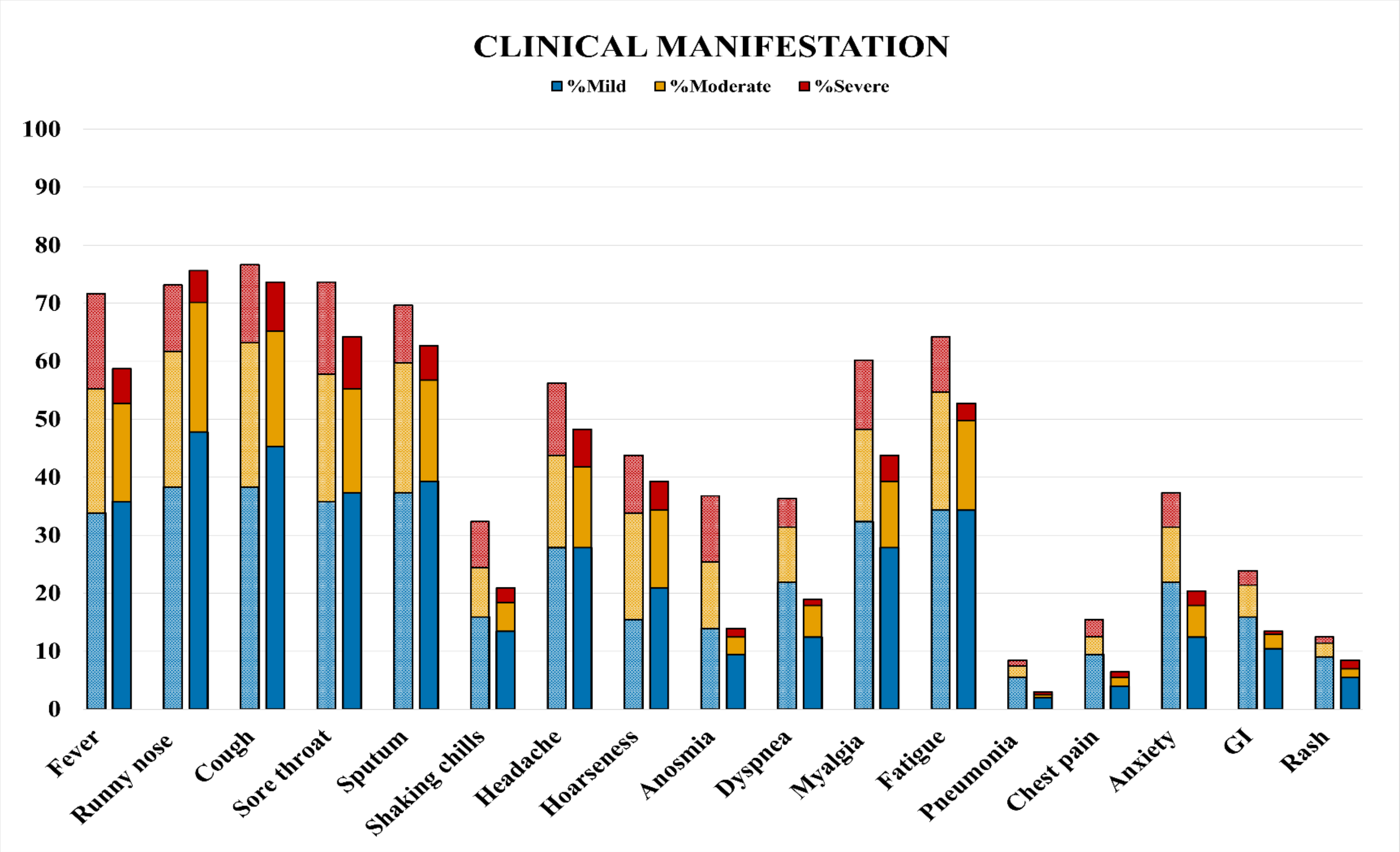
